# Pan-Cancer Proteogenomic Network Analysis Uncovers Immune Microenvironment and Prognostic Biomarkers

**DOI:** 10.1101/2025.01.15.24319471

**Authors:** Wenhan Shi, Runing Guan, Jiafeng Li, Dongjing Chen

**Author notes:** Correspondence: Prof. Dongjing Chen, AmazingX Academy, Headquarters Center, Nanshan District, Shenzhen.

## Abstract

Pan-cancer analysis provides valuable insights into common and distinct molecular mechanisms across various cancer types. Co-expression networks, which captured the cordinated expression of genes and proteins, offer potentials for identifying robust biomarkers and targeted therapies. However, current-existing co-expression construction method involves either RNA or Protein data in isolation, limiting our understanding on multi-perspective interactions in cancer. To address this gap, we integerated both the RNA and Protein data to constrcut a comprehensive pan-cancer co-expression network, combining both gene and proteomic layers of information across cancer types. This approach enables the identification of previously undetected co-expression relationships, highlighting key regulatory patterns in cancer analysis. Our study constructed a multi-perspective co-expression network, after that, we identified several conservation modules and compare the modules identified from RNA and Protein. We found out that the conservation modules identified differ significantly from RNA and Protein data. However, there are certain shared conservation modules, especially those related to IFN-gamma, IFN-alpha, and the activation of JAK_STAT. Then we identified several important immune-related pathways using RNA and Protein data and uncovered some modules with high prognostic value across cancers. As a novel pan-cancer analysis approach, it holds promise for advancing future cancer research, from the discovery of novel biomarkers to the design of personalized cancer therapies, thereby enabling more thorough and holistic cancer analysis.

## Introduction

As one of the most complex, dynamic human diseases, cancer’s complexity stems from its multifaceted nature, encompassing genetic, epigenetic, and environmental factors. At the genetic level, cancer involves mutations in a wide range of genes, leading to diverse and unpredictable responses in patients.[1] Additionally, the interaction between cancer cells and the microenvironment (TME) complicates the situation [2] by dysregulating the cell-cell interaction pathways. Epigenetic modifications also play a crucial role, modifying the gene expression without changing the DNA sequence[3] and contributing to the unpredictability of tumors. This intricate network of factors makes cancer a highly complex and heterogeneous disease, challenging both diagnosis and treatment. Meanwhile, biological network analysis offers significant advantages in tumor research by providing a systematic view of the complex interactions [4]within the biological system. By mapping out networks of genes, proteins, and other molecules, researchers can identify crucial nodes and pathways involved in cancer and gain insight into how disruptions in one part of the network [5]might impact other components. Therefore, leveraging systematic biological network analysis in pan-cancer research allows for a more comprehensive and holistic approach to tackling this complex disease.

According to the existing experimental conclusions, most studies have focused on building the co-expression network based on RNA-seq data. However, recent advancements in proteomics have revealed that the correlation of mRNA and protein is often lower than anticipated[6] due to factors like translation and transcription modifications. Therefore, more than building a co-expression network relying entirely on RNA-seq data is required for a comprehensive and targeted tumor analysis. Fortunately, the newly developed database provides us with an extensive range of proteome information, allowing us to build a more precise co-expression network based on the proteome data to capture a more accurate representation of biological processes and interactions. [7]

Prognostic analysis is crucial for predicting disease outcomes, guiding treatment decisions, and improving patient management. [8]Accurate prognostic assessments can enhance the effectiveness of medical intervention and increase individual patients’ particularizability[9]. Integrating network analysis into prognosis research provides a significant advantage by uncovering complex interactions among genes, proteins, and other molecules. By constructing and analyzing the co-expression network, researchers can identify vital prognostic components in the network and their roles in regulating specific mechanisms in cancer. Currently, no such research about networks based on large-scale proteome data and their corresponding prognosis is available for applications in cancer analysis. Therefore, in our study, we focus on building a precise proteome-based co-expression network that helps with prognosis analysis, and we believe that our study will not only improve the precision of prognostic prediction[10] but also reveal potential targets for therapeutic interventions.

The modules within biological networks are often linked to specific phenotypes, making them valuable tools for understanding the underlying mechanisms of diverse disease cases.[11] Researchers can uncover the connection between network modules and certain phenotype expressions by systematically searching for different phenotypes, such as the immune cell abundance. This approach is crucial for identifying critical regulatory pathways and potential therapeutic targets, ultimately leading to a more comprehensive understanding of disease causation and effective treatment strategies.

Therefore, our research focuses on constructing a cancer co-expression network based on large-scale proteome data, providing a more comprehensive understanding of the molecular interactions involved in cancer. We holistically describe co-expression networks and prognostic modules in cancer and reveal multiple regulatory mechanisms related to cancer prognosis.

Further, our network serves as a valuable tool for evaluating the conservation of these modules and genes across different cancer types. It enlightens researchers with the underlying conservation patterns shared by different tumors. Ultimately, our work can potentially guide future therapeutic strategies by revealing conserved biological processes that influence tumor behavior across the cancer spectrum.

## Method

### Clinical and molecular data sets

The proteomics data used in this study were obtained from the Clinical Proteomic Tumor Analysis Consortium (CPTAC), which has characterized over 1,000 treatment-naïve primary tumors spanning 10 different cancer types. The cancer types included in the CPTAC dataset are breast cancer (BRCA), ovarian cancer (OV), colon cancer (COAD), clear cell renal cell carcinoma (ccRCC), uterine corpus endometrial carcinoma (UCEC), lung adenocarcinoma (LUAD), liver hepatocellular carcinoma (LIHC), glioblastoma multiforme (GBM), pediatric brain tumors (PBT), and pancreatic cancer (PAAD). In total, we collected proteomic, RNA, and phenotypic data from 1,043 cancer patients across these 10 cancer types.

For proteomics data, mass spectrometry-based proteomic profiling was employed, using label-free quantification to measure protein expression levels. The data were further processed with normalization and batch correction to remove technical variability, ensuring accurate cross-sample comparisons. Specifically, we downloaded RNA-seq data at the gene level, processed using the RSEM (RNA-Seq by Expectation-Maximization) algorithm, ensuring high-quality and reliable quantification of gene expression.

The dataset covers a comprehensive range of phenotypes, including clinical factors such as age, BMI, sex, histologic grade, tumor stage, overall survival, progression-free survival, and tumor size, offering insight into patient prognosis. Immune phenotypes are analyzed using tools like CIBERSORT and xCell, providing estimates of various immune cells (e.g., B cells, T cells, macrophages, NK cells, neutrophils, monocytes) within the tumor microenvironment. Additionally, hallmark pathways (e.g., inflammatory responses, hypoxia) and mutation signatures (e.g., SBS1, APOBEC) contribute to understanding the molecular mechanisms of cancer progression, supported by ImmuneScore and StromalScore data on tumor-immune interactions.

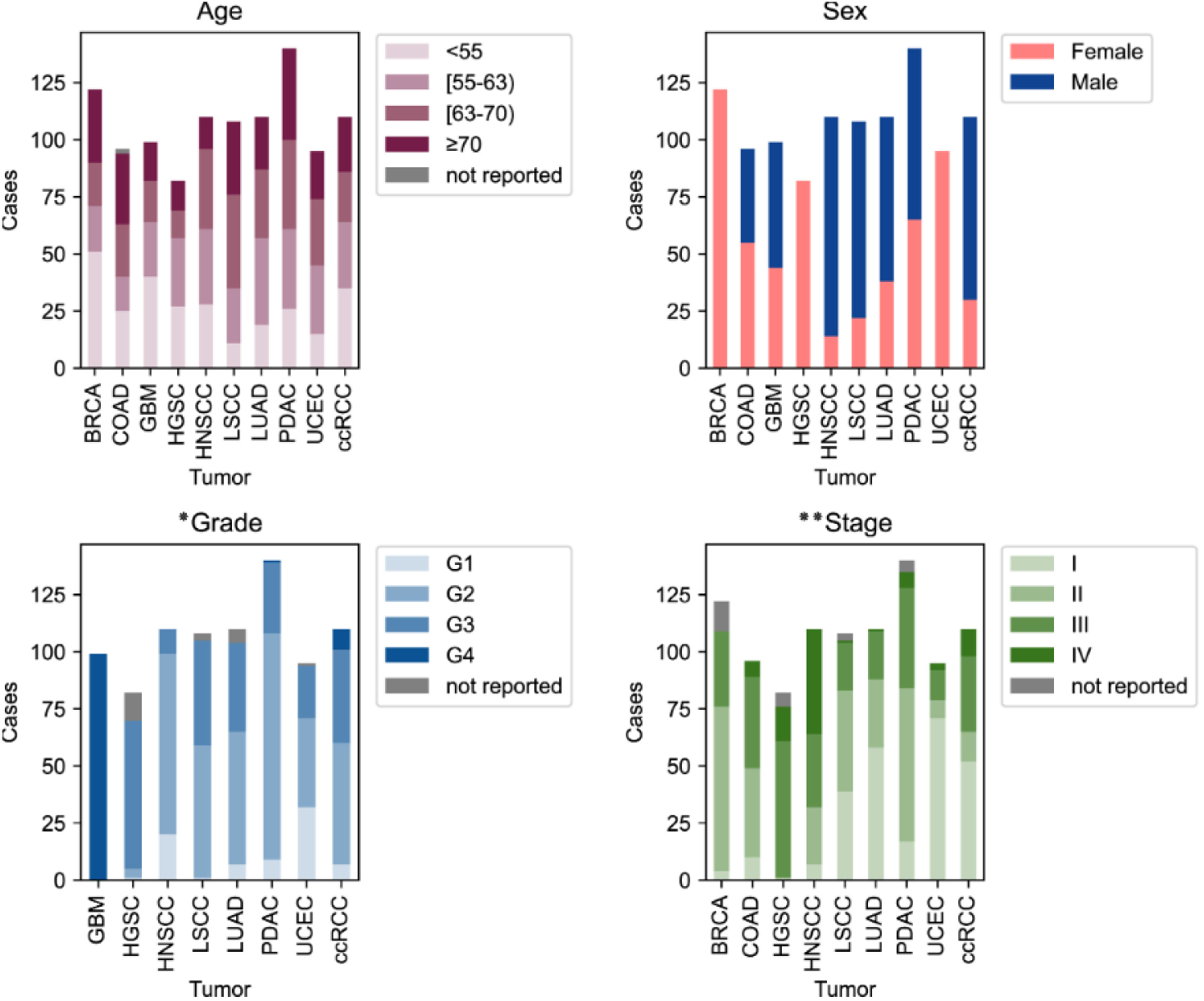

### Co-expression network construction

The co-expression network was constructed using the WGCNA package in R(4.4.1), which is a systematic software package for analyzing and constructing gene networks across diverse state or condition samples. The following steps outline the process of network construction with detailed parameter settings:

We utilized the WGCNA package in R to construct a gene co-expression network from normalized log-transformed gene expression and protein data. The analysis commenced with data preparation, where gene identifiers were truncated, and gene symbols were integrated through annotation.

Subsequently, genes were filtered based on their median absolute deviation (MAD), retaining those with MAD greater than the 50th percentile variability. Samples and genes with excessive missing values were excluded based on the output from goodSamplesGenes.

The network construction involved determining a soft thresholding power using the scale-free topology model fit, targeting an R^2 value of 0.85 for optimal network representation. We employed the “unsigned” network type, allowing for the consideration of both positive and negative correlations. Module detection was performed using the blockwiseModules function with a merge cut height of 0.25, ensuring the identification of distinct gene co-expression modules. The minimum module size was set to 30 genes to maintain biological relevance.

For visualization, module colors were assigned, and a heatmap of the gene dendrogram was created. Module eigengenes (MEs) were calculated and used to generate an adjacency heatmap to explore module relationships. The network heatmap was constructed, highlighting gene-to-gene connections with enhanced visibility for moderately strong links through power transformation.

Phenotypic data integration allowed us to correlate module eigengenes with clinical traits using either Pearson correlation. The resulting correlations were visualized, and significant associations were identified. Genes were further analyzed for their association with phenotypic traits, pinpointing those with high module membership and significant trait correlation, potentially highlighting key genes of biological importance.

### Identify conserved modules

To explore the conservation between modules, we introduced the Jaccard index for assessment. This involved comparing the genes within modules of one cancer type to the network modules present in other cancer types. The similarity between each pair of modules was quantified using the Jaccard index, which is calculated based on the genes from the respective modules, denoted as set A and set B. To investigate the conservation between modules, we employed the Jaccard index for assessment. Initially, we evaluated the conservation of modules identified from RNA and protein data within the same cancer type. Subsequently, we computed the pairwise module similarity across all cancer types for networks constructed from protein data and RNA data, respectively. Finally, we identified clusters of highly conserved modules through hierarchical clustering

### Module annotation and prognosis analysis

To understand the functions affected by genes within each module, we performed functional annotation of the modules. On one hand, we calculated the relationship between module eigengenes and phenotypes using WGCNA. On the other hand, we conducted enrichment analysis using the clusterProfiler R package, based on GO, KEGG, and Reactome databases. We corrected the p-values for FDR and retained only those terms with FDR < 0.05 and ranked in the top 5 within each database. To assess the relationship between modules and prognosis, we performed survival analysis for each module. We obtained the batch effects normalized mRNA data and survival data from the TCGA Pan-Cancer (PANCAN) database. For each module, we conducted PCA on all the genes and extracted PC1 to use as a grouping criterion, dividing the samples into PC1 high-expression and PC1 low-expression groups. To test whether the expressions of module eigengenes significantly affect survival time (P < 0.05), we built a Cox proportional-hazard model using the R function _“_coxph._”_ The hazard ratio reflects how expression levels influence the rate of patient survival, where an increase in the death hazard corresponds to a decrease in survival length. Additionally, we conducted the same analysis considering all cancer samples as a whole.

## Result

### Pan-cancer co-expression networks and modules

We performed WGCNA analysis on ten types of cancer based on RNA data and protein data, constructing ten protein co-expression networks and ten RNA co-expression networks. To identify key gene interactions, we retained only the top 50% of genes by median absolute deviation (MAD) from the protein data for network analysis. Through hierarchical clustering, numerous modules were identified from each network, with specific results presented in Table 1.

**Table 1:** The number of genes used to construct the pan-cancer network and the number of modules identified.

| Cancer | Protein |  | RNA |  |
| --- | --- | --- | --- | --- |
|  | Number of genes | Number of modules | Number of genes | Number of modules |
| BRCA | 4440 | 13 | 4436 | 12 |
| CCRCC | 4056 | 13 | 4052 | 9 |
| COAD | 3173 | 13 | 3166 | 11 |
| GBM | 2916 | 9 | 2916 | 7 |
| HNSCC | 4269 | 14 | 4267 | 10 |
| LSCC | 4985 | 13 | 4983 | 9 |
| LUAD | 4682 | 18 | 4680 | 13 |
| OV | 3823 | 14 | 3819 | 10 |
| PDAC | 3996 | 11 | 3994 | 11 |
| UCEC | 4479 | 12 | 4476 | 9 |

The scientific question that initially concerned us was whether there are differences between networks constructed based on protein data and RNA data. Therefore, we assessed the conservation of modules identified by networks built from different data types within the same type of cancer using the Jaccard index. It is noteworthy that we used the same set of genes to construct networks of different data types. The results showed that the modules identified by networks built from different data types were extremely non-conservative, with an average Jaccard index of 0.04 and the highest Jaccard index was 0.65(Figure 1A).

**Figure 1:**
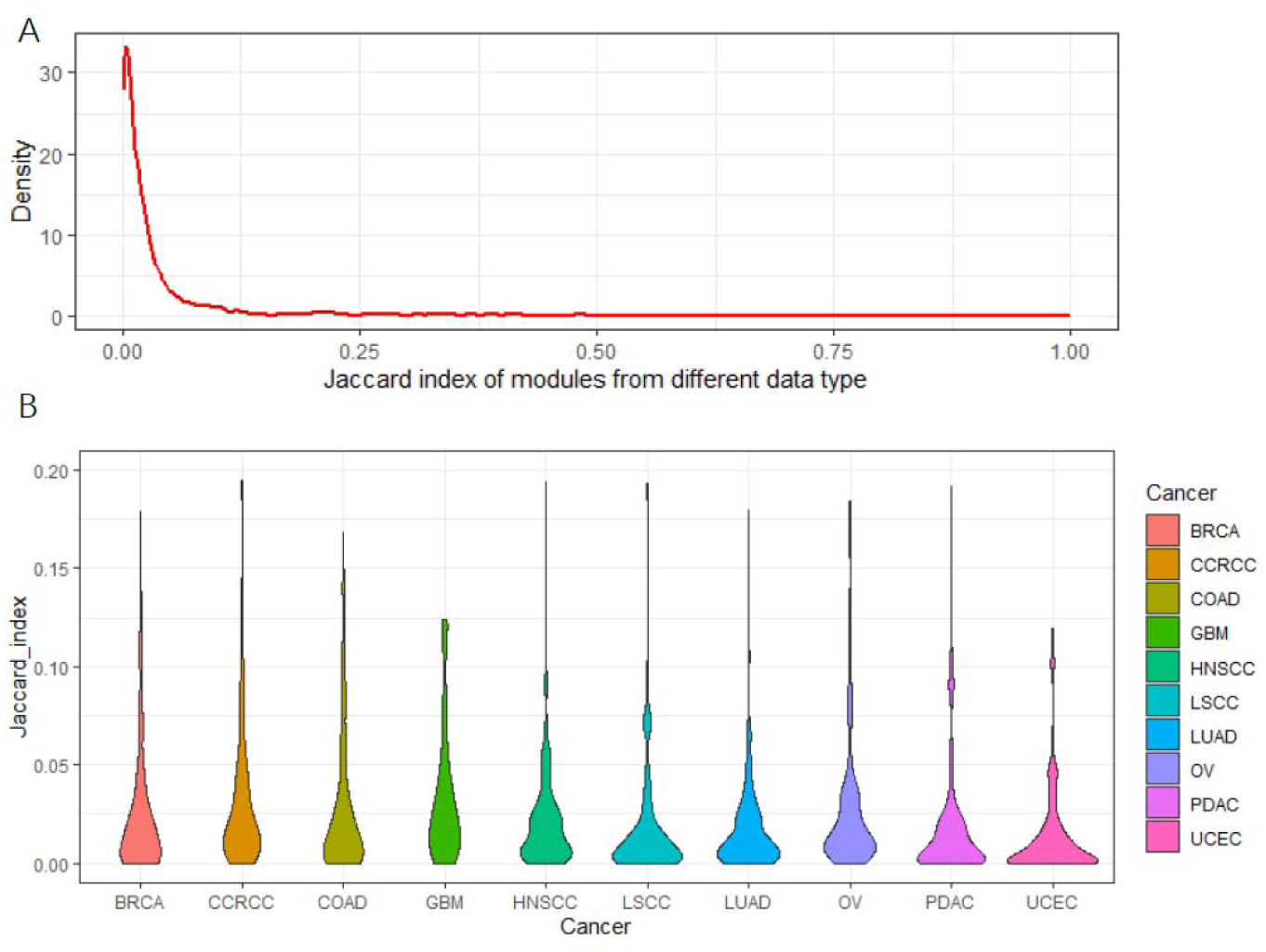
The distribution of Jaccard Index between modules constructed based on protein data and RNA data.

Moreover, this non-conservatism is shared across different types of cancer(Figure 1B). This suggests that when studying biological processes through interaction networks, multiple data sources must be considered simultaneously.

### Identify pan-cancer preserved modules

We did the preserved analysis of proteins in between modules and different cancers (Fig. 2A) and found the conserved part in pan-cancer. Among them, cluster 1 (Fig. 2B) contains 9 cancers—HNSCC, PDAC, UCEC, CCRCC, LUAD, COAD, GBM, BRCA, and LSCC; and cluster 2 (Fig. 2C) contains 6 cancers—LSCC, BRCA, HNSCC, COAD, LUAD, and OV. Among them, the main function of the cluster module is focused on the final culmination in the formation of thrombin, the enzyme responsible for the conversion of soluble fibrinogen to the insoluble fibrin clot[11]; and for cluster 2, it is mainly focused on defense response to the virus, which is the reactions[12] triggered in response to the presence of a virus that acts to protect the cell or organism.

**Figure.2:**
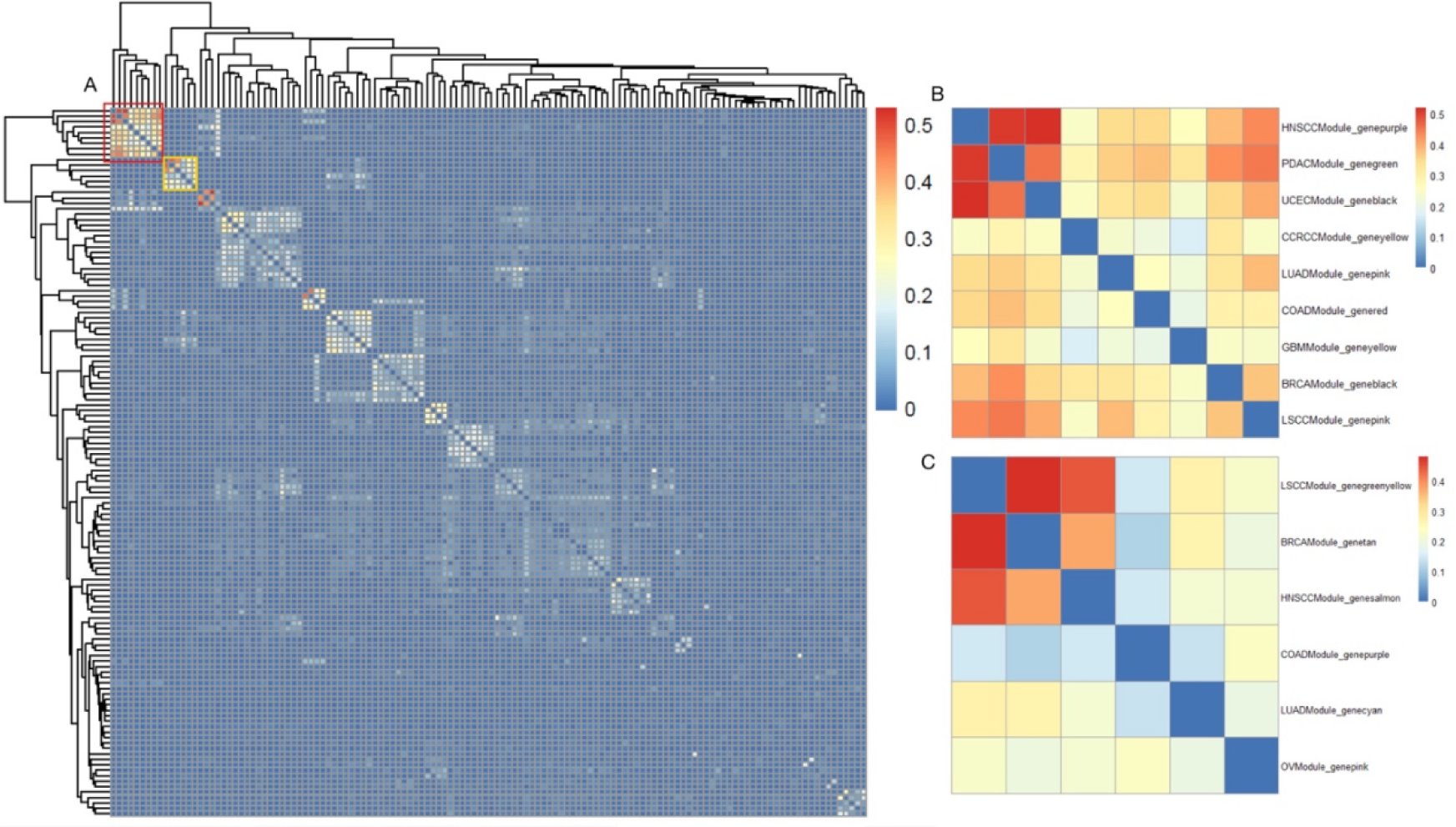
Conservation analysis of proteins in between modules of different cancers. (A) Conservation analysis of proteins in all modules of different cancers, where the color intensity represents the conservation level on the scale of 0 to 0.5; the red block refers to Figure 2B and the yellow block refers to Figure 2C. (B) Conservation analysis of proteins of different modules in 9 of the cancers. Rows represent the various modules in 9 cancers, and the color intensity represents the conservation level on the scale of 0 to 0.5. (C) Conservation analysis of proteins of different modules in 6 of the cancers. Rows represent the different modules in those 6 cancers, and the color intensity represents the conservation level on the scale of 0 to 0.5.

We did the preserved analysis of proteins in between modules and different cancers (Fig. 3A) and found the conserved part in pan-cancer. Among them, cluster 1 (Fig. 3B) contains 9 cancers—PDAC, UCEC, LSCC, HNSCC, OV, CCRCC, COAD, and LUAD; and cluster 2 (Fig. 3C) contains 8 cancers—UCEC, LSCC, GBM, OV, PDAC, BRCA, HNSCC, and LUAD. In regard to cluster 1, the function of the modules primarily concentrates on immunoglobulin production, which allows the immune system to recognize and effectively respond to a myriad of pathogens[13]; cluster 2, similar to cluster 2 in protein, focuses on the function of defense response to the virus.

**Figure 3:**
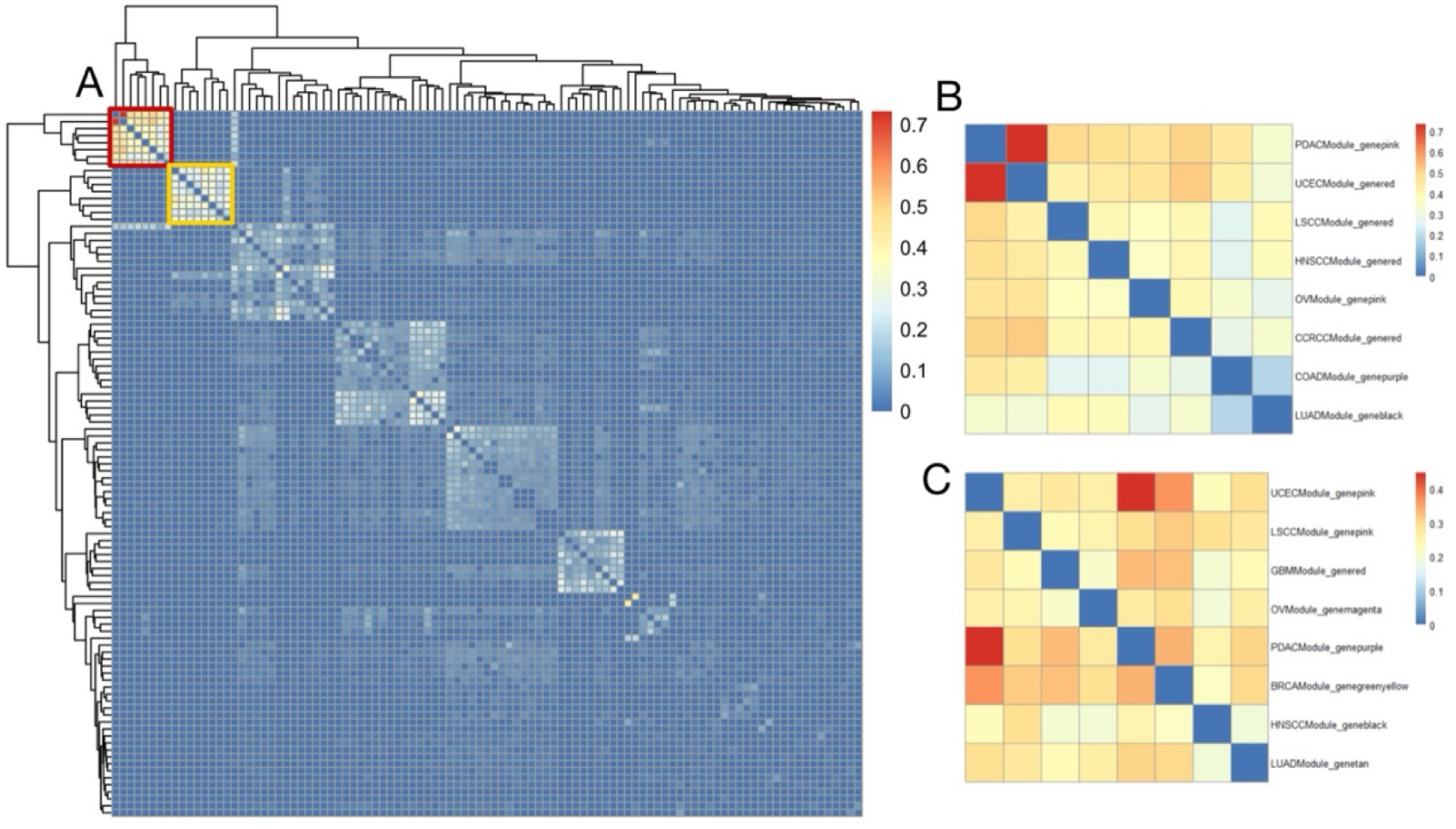
Conservation analysis of RNAs in between modules of different cancers. (A) Conservation analysis of RNAs in all modules of different cancers, where the color intensity represents the conservation level on the scale of 0 to 0.7; the red block refers to Figure 3B and the yellow block refers to Figure3C. (B) Conservation analysis of RNAs of different modules in 8 of the cancers. Rows represent the various modules in 8 cancers, and the color intensity represents the conservation level on the scale of 0 to 0.7. (C) Conservation analysis of RNAs in different modules in 8 of the cancers. Rows represent the different modules in those 8 cancers, and the color intensity represents the conservation level on the scale of 0 to 0.4.

### Functions Impacted by Preserved Pan-Cancer Modules

As shown in Figure 4A Protein cluster 1 can be identified that hypoxia is highly related to several protein clusters, with the highest average relationship value of 0.48. Here, hypoxia represents a condition where tissues experience low oxygen levels, and it drives several processes that promote cancer progression in the tumor microenvironment. This is mainly because hypoxia promotes abnormal, yet excessive, blood vessel formation and triggers the epithelial-to-mesenchymal transition (EMT) phenotype, which enhances cell movement and metastasis. Additionally, hypoxia modifies cancer cell metabolism and supports therapy resistance by inducing a quiescence state in the cell. [12] As part of the cause of hypoxia, the epithelial-to-mesenchymal transition is the second highly related phenotype to the protein clusters, with an average relationship value of 0.45. This is mainly because EMT can enhance the ability of cells to invade the surrounding tissues. [13] Other phenotypes with relatively high relationship scores are myogenesis, KRAS signaling up, stromal score, and the activation of the TGFb pathway, all with a relationship score above 0.4. According to the data of Protein cluster 2 (Fig 4B), we can conclude that the activation of the JAK-STAT pathway has the highest relationship with the protein clusters, with an average relationship value of 0.89. This phenomenon is primarily because the activation of the JAK_STAT pathway often drives cancer cell proliferation and metastatic behaviors. [14]The phenotype with the second highest relationship value is the dysfunction of interferon-alpha-response (IFN-alpha), with a relationship score of 0.86. The IFN-alpha can partly cause tumors because it induces cell apoptosis. [15] Other phenotypes closely related to the clusters in protein 2 are INF-gamma, JAK-STAT signaling, allograft rejection, inflammatory response, complement, macrophage-M1, immune score, and myeloid dendritic cell activated, all with a relationship score above 0.5.

**Figure 4:**
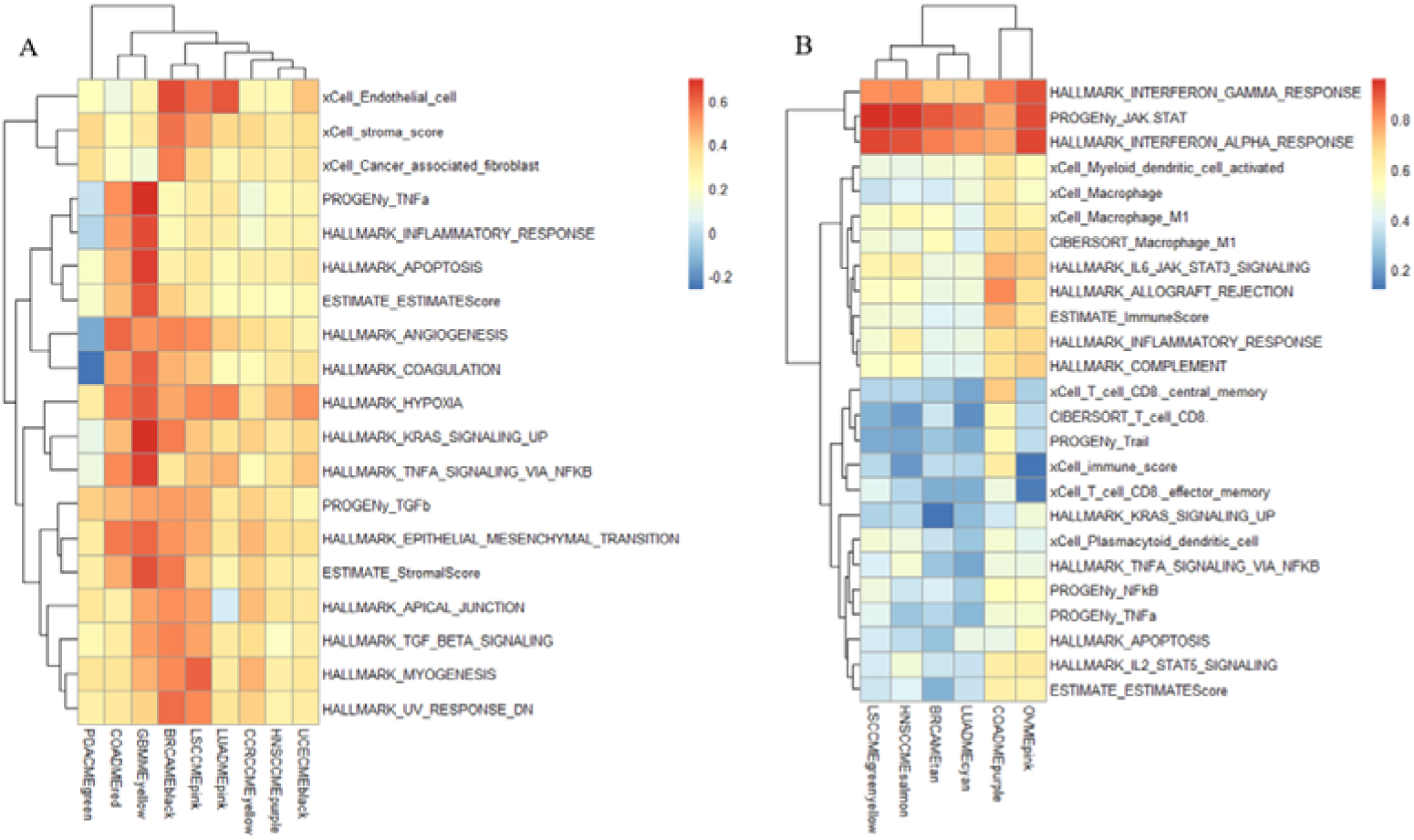
Characterization of conserved module clusters based on protein data. (A) Biological functions of protein cluster 1. (B) Biological functions of protein cluster 2.

Moving on to the RNA data, we can conclude that in RNA cluster 1(Figure 5 A), the phenotype with the highest relationship with the clusters is the B cell plasma, with a relationship score of 0.61. Recent data shows that B cell plasma is located in tumors, and the dysfunction of the B cell plasma can produce specific antibodies that drive distinct immune responses. [16]Another noticeable phenotype shown in RNA 1 is the immune score, with a relationship score of 0.59. This is relatively understandable since the cancer cells can reduce the expression of molecules (like MHC class I proteins) that present abnormal proteins (antigens) to immune cells, making it harder for T-cells to recognize them as dangerous.

**Figure 5:**
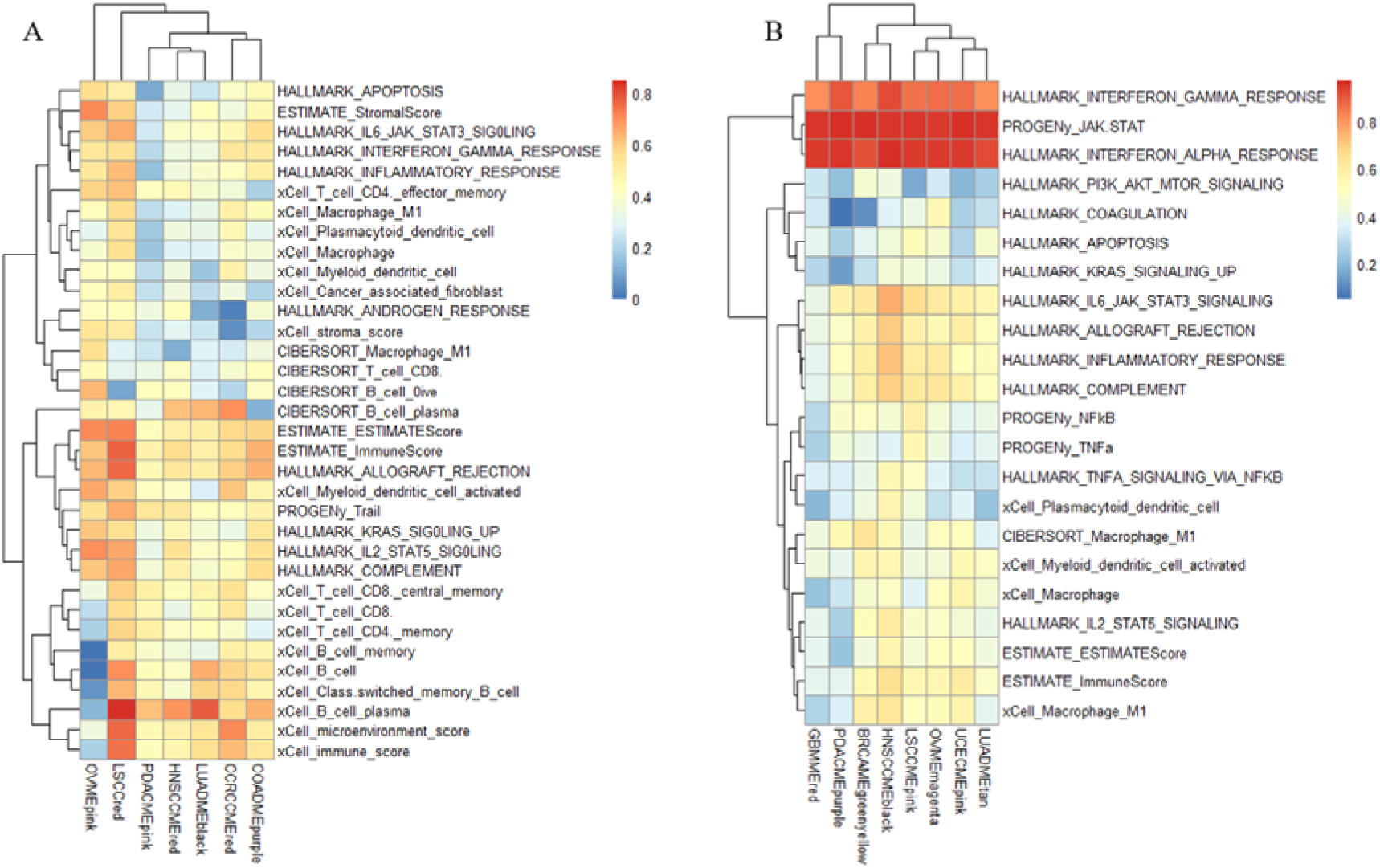
Characterization of conserved module clusters based on RNA data. (A) Biological functions of protein cluster 1. (B) Biological functions of protein cluster 2.

Other phenotypes with relatively high correlation with the RNA clusters are allograft rejection, microenvironment score, Il2-STAT5-signaling, trail, and complement, all with a correlation value above 0.5. Similarly, according to the data in RNA cluster 2(Figure 5 B), it can be identified that the activation of the JAK_STAT pathway and INF-alpha are the two phenotypes with the highest correlation to the clusters, ranking 0.95 and 0.86 respectively. Other phenotypes with relatively high correlation are IFN-gamma, IL6_JAK_STAT3_signaling, inflammatory response, complement, allograft rejection, and immune score, all with a correlation score above 0.5.

Now, it is shown that the network based on the protein and RNA databases shares certain similarities. As shown in both the graph of protein2 and RNA2, both recognize that IFN-alpha, IFN-gamma, and the activation of JAK_STAT are of high correlation with the clusters, indicating that these are the shared characteristics of cancer. However, there are also significant differences between the network based on RNA and protein. In protein 1, hypoxia is highly signified as a typical phenotype shared by many cancer modules, such as the GBM (MEyellow), LUAD (MEpink), and COAD (MEred). Meanwhile, RNA1 showcases nothing about these module clusters, and hypoxia isn’t highlighted as a representative phenotype shared by the modules. Similarly, the EMP is also a phenotype shared between the cancer modules according to protein 1, yet no evidence is shown in RNA1. The same thing happens to the myogenesis and KRAS signal. Yet the RNA data also reveals some phenotypes shared between the modules and not recognized by the protein 1 data. In RNA 1, the B cell plasma is highly correlated to the modules. Still, in protein 1, this phenotype only has a relationship score of −0.02, signifying the differences in the analysis result from the two data. A similar situation also falls on the phenotypes of allograft rejection, microenvironment score, IL2_TAT5 signaling, and trail, where these phenotypes are only recognized as shared phenotypes in RNA1 but no related signs in protein1.

The data can also be used to analyze the group clusters of these cancer modules. From the graph of protein1, we can conclude that COAD (MEred), GBMM (MEyellow), BRCA (MEblack), and LSCC (MEpink) can be divided into the same group, exhibiting a relatively high correlation with almost all the phenotypes shown in the graph. In contrast, LUAD (MEpink), CCRCC (MEyellow), HNSCC (MEpurple), and UCEC (MEblack) can be divided into another group, exhibiting a relatively low correlation with the phenotypes compared to the previous group. From the graph of protein2 provided, we can conclude that the LSCC (MEgreenyellow), HNSCC (MEsalmon), BRCA (MEtan), and LUAD (MEcyan) can be divided in the same group, exhibiting little correlation with the last eight phenotypes shown in the graph; meanwhile, COAD (MEpurple) and OV (MEpink) can be divided in another group, exhibiting certain level of correlation with the eight phenotypes at the back shown in the graph. From the graph of RNA1, we can divide the group by putting LSCC (red) separately in one group. In contrast, the rest of the modules are divided into another group since they aren’t correlated to every phenotype shown in the graph. Still, the LSCC (red) module showcases a certain level of correlation with every phenotype shown in the graph. From the graph of RNA2, all the modules exhibit relatively similar correlations with the phenotypes, with high relationships between IFN-alpha and IFN-gamma and the activation of JAk_STAT. Therefore, it is better to put all the modules in RNA2 in the same group because of their resemblance in the correlation pattern.

### The prognostic features of conserved module and immune related module

To elucidate the prognostic patterns of the identified modules, an extensive investigation was conducted to evaluate their prognostic significance across various cancers. The analysis focused on assessing the differential prognostic value of these modules, revealing four with particularly high prognostic relevance based on RNA and protein data.

According to the protein data, the BRCA (MEpink) (Fig.4A) and COAD (MEyellow) (Fig.4B) modules exhibited strong prognostic value across 15 different cancer types, as shown in the forest plots below. This suggests that these modules may play key roles in predicting disease outcomes in a wide range of cancers. Meanwhile, based on the RNA data, the BRCA (MEquoiseCox) (Fig.4C) and LUAD (MEbrown) (Fig.4D) modules emerged as highly prognostic across 16 cancers, further highlighting their potential as important biomarkers for cancer prognosis, as also depicted in the accompanying forest plots.

**Fig. 6:**
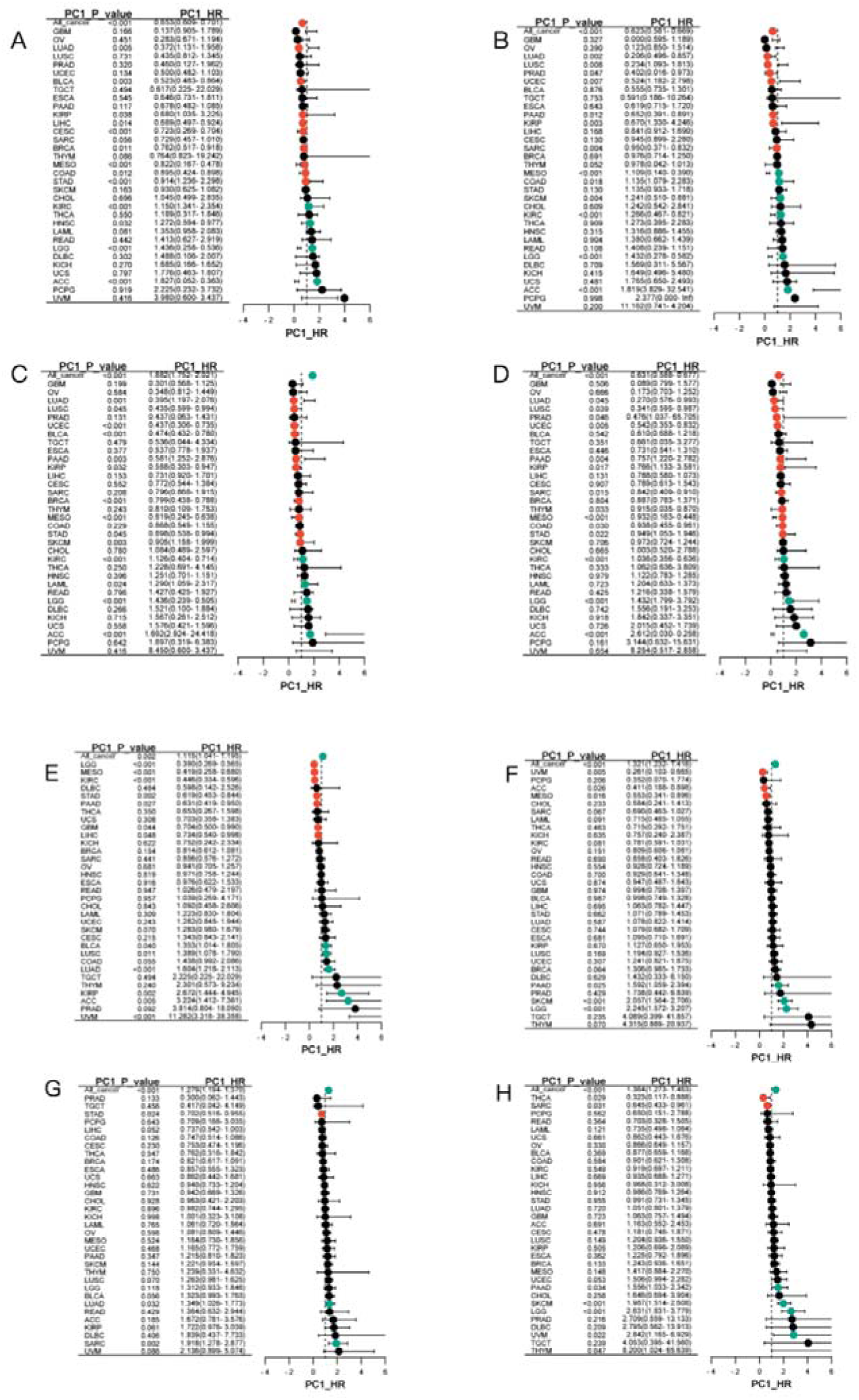
Forest figures presenting the prognostic functions of the modules. The figures displayed are with a 95% condifence interval of the result. (A) is the forest figure for BRCA(MEpink), (B) is the forest figure for COAD(MEyellow), figure 5(C) is the forest figure for BRCA(MEquoiseCox) and (D) is the forest figure for LUAD(MEbrown). (E) the forest graph showing the prognostic value of the protein cluster1, the red module of the cancer COAD. (F) the forest graph showing the prognostic value of the protein cluster2, brown module of the cancer OV. (G) the forest graph showing the prognostic value of the RNA cluster1, the pink module of the cancer OV. (H) the forest graph showing the prognostic value of RNA cluster2, the greenyellow module of the cancer BRCA. (I) the forest graph showing the protein immune related red module of the cancer GBM. (J) the forest graph showing the RNA immune related brown module of the cancer OV.

In addition to the between-group analysis for the modules, we also conducted a within-group analysis by focusing on the HR values and drawing a forest plot. In the protein analysis, we found that COADMEred (Fig. 4E) and OVMEpink (Fig. 4F), belonging to cluster1 and cluster2 respectively, have strong prognostic values as shown in the following figures. Meanwhile, in RNA, OVMEpink (Fig. 4G) and BRCAMEgreenyellow (Fig. 4H), belonging to cluster1 and cluster2 respectively, also performed strong prognostic values.

## Discussion

Our research offers a comprehensive exploration of the pan-cancer co-expression network using both the RNA and the Protein data, leading to significant findings on module conservation and prognostic implications. By constructing co-expression networks, we were able to identify several distinct modules that reflect the underlying biological processes in cancer. One key finding is the highlighted difference for the distinct modules identified from RNA and protein within the same cancer types, emphasizing the importance of analyzing both networks when conducting therapeutic research on cancer.

In terms of module conservation, although our analysis revealed remarkable differences between modules, there are certain conserved modules at the pan-cancer level, particularly those related to IFN-gamma, IFN-alpha, and the activation of JAK_STAT. Furthermore, our evaluation of immune-related modules underscores the importance of immune pathways in cancers. Several modules were found to be related to immune pathways, such as the BRCA(MEred), COAD(MEpurple), CCRCC(MEgreen), GBM(MEblue), LSCC(MEturquoise), and HNSCC(MEblue). The finding of these immune pathways reflect the significant role of the immune microenvironment in shaping the cancer outcomes.

Finally, the research delved into the relationship between these identified modules and cancer prognosis. We uncovered several prognostic modules using both the RNA and Protein data, such as the BRCA(MEpink), COAD(MEyellow), BRCA(MEquoisecox), and COAD(MEyellow) identified in Result 4. Interestingly, some of the conserved and immune-related modules also demonstrated prognosis relevance, suggesting that these modules might not only uncover the biological processes behind the cancers but also predict future outcomes for the patients.

Our findings mark a significant departure from conventional analysis methods, where a single data type, especially the RNA data, is used to infer the gene and protein interaction in network analysis. By incorporating both the RNA and the Protein data derived from the same sample, our study uncovers the complexity and unpredictability of cancer co-expression networks. The significant differences between modules identified by RNA and Protein suggest that these two aspects capture distinct biological processes. While RNA provides a snapshot of gene expression level, Protein reflects the modification during translation and transcription, signifying cancer network dynamics.

The differences observed with RNA and Protein data also signify the limitation of using solely one type of data for network construction. RNA-based networks, while reflecting the cancer characteristics at the gene expression level, might misidentify the processes undergoing on the protein level. Similarly, protein-based networks can accurately reflect the processes happening on the protein level but might overlook the transcriptional modification captured in RNA data. Therefore, our study underscores the necessity of integrating multi-perspective sources to construct more comprehensive and representative interactive networks.

Building on the findings of RNA and protein-based network analysis, this study’s future applications are vast, particularly in personalized medicine and targeted cancer therapies. The identified conservative, immune-related, and prognostic modules are newly discovered and can be potential targets for future cancer research. This dual-perspective research can be instrumental in finding novel biological markers that are not detected by RNA or Protein data alone, providing a more comprehensive view of pan-cancer analysis.

Specifically, some of the potential applications of this approach can be predicting patient-specific responses to certain therapies and promote the development of combined therapies toward interaction between immune and cancer cells, both in RNA level and transcriptional level. By using both data types, clinicians could design personalized treatment plans that target not only the gene expression but also the proteomic activities for cancer patients.

In conclusion, the integration of RNA and protein-based pan-cancer network construction provides a powerful tool for addressing complex challenges in cancer treatment. It has the potential to guide future cancer analyses from novel biomarkers to personalized cancer therapies, paving the way for more comprehensive and accurate cancer analysis.

## Data Availability

All data produced are available online at https://portal.gdc.cancer.gov/

## Acknowledgement

In one of our elective courses: synthesis biology, when our instrcutor Jia You was explaining the process of the formation of protein, she mentioned that even though RNA serves as the guide for protein, there can exist unpredictable modification in the folding of protein.

Therefore, the result based on RNA data and Protein data can sometimes be distinct in complex studies. Thinking of the most dynamic and complex biological challenge currently in the world, cancer analysis is undoubtly worth noticing. We then came up with the idea of doing cancer analysis using both the RNA and Protein data, trying to uncover some previously undected discoveries. After many intense discussions with our instructor, Ms. You advised us to consider biological challenges systematically, instead of solely understand each individual molecule which might lead to interpretation biases such as overlooking the significance of a single gene. In terms of cancer, a biological issue involving multiple aspects and con’t be easily divided into a single column, we eventually decided to construct a co-expression network to uncover the complex interactions between different modules.

In our study, Wenhan Shi is mainly in charge of formal analysis, programming, paper-searching, writing–original draft, writing–review and editing; while Runing Guan is in charge of data collection, programming, paper-searching, writing–original draft, writing–review and editing.

We would like to deliever our sincere thankfulness to our instructor Jia You, the teacher of biology and the elective course of synthesis biology, for her kind advises and instructions on topic choosing. She didn’t receive any reward from the author.

We would also like to express our thank to the Sponsors of Qiu Chengtong academic competition, for they provided us with a broader stage to share our research and we’ve learned a lot about biological tecnology throughout our research.

At last, we would like to express our sincere gratefulness again to everyone who supported and helped us.

